# Epidemiological and clinical characteristics of the early phase of the COVID-19 epidemic in Brazil

**DOI:** 10.1101/2020.04.25.20077396

**Authors:** William Marciel de Souza, Lewis Fletcher Buss, Darlan da Silva Candido, Jean-Paul Carrera, Sabrina Li, Alexander E. Zarebski, Maria F. Vincenti-Gonzalez, Janey Messina, Flavia Cristina da Silva Sales, Pamela dos Santos Andrade, Carlos A. Prete, Vítor Heloiz Nascimento, Fabio Ghilardi, Rafael Henrique Moraes Pereira, Andreza Aruska de Souza Santos, Leandro Abade, Bernardo Gutierrez, Moritz U. G. Kraemer, Renato Santana Aguiar, Neal Alexander, Philippe Mayaud, Oliver J. Brady, Izabel Oliva Marcilio de Souza, Nelson Gouveia, Guangdi Li, Adriana Tami, Silvano Barbosa de Oliveira, Victor Bertollo Gomes Porto, Fabiana Ganem, Walquiria Aparecida Ferreira de Almeida, Francieli Fontana Sutile Tardetti Fantinato, Eduardo Marques Macário, Wanderson Kleber de Oliveira, Oliver G. Pybus, Chieh-Hsi Wu, Julio Croda, Ester C. Sabino, Nuno Rodrigues Faria

## Abstract

**Background:** The first case of COVID-19 was detected in Brazil on February 25, 2020. We report the epidemiological, demographic, and clinical findings for confirmed COVID-19 cases during the first month of the epidemic in Brazil.

**Methods:** Individual-level and aggregated COVID-19 data were analysed to investigate demographic profiles, socioeconomic drivers and age-sex structure of COVID-19 tested cases. Basic reproduction numbers (*R*_*0*_) were investigated for São Paulo and Rio de Janeiro. Multivariate logistic regression analyses were used to identify symptoms associated with confirmed cases and risk factors associated with hospitalization. Laboratory diagnosis for eight respiratory viruses were obtained for 2,429 cases.

**Findings:** By March 25, 1,468 confirmed cases were notified in Brazil, of whom 10% (147 of 1,468) were hospitalised. Of the cases acquired locally (77·8%), two thirds (66·9% of 5,746) were confirmed in private laboratories. Overall, positive association between higher per capita income and COVID-19 diagnosis was identified. The median age of detected cases was 39 years (IQR 30-53). The median *R*_*0*_ was 2·9 for São Paulo and Rio de Janeiro. Cardiovascular disease/hypertension were associated with hospitalization. Co-circulation of six respiratory viruses, including influenza A and B and human rhinovirus was detected in low levels.

**Interpretation:** Socioeconomic disparity determines access to SARS-CoV-2 testing in Brazil. The lower median age of infection and hospitalization compared to other countries is expected due to a younger population structure. Enhanced surveillance of respiratory pathogens across socioeconomic statuses is essential to better understand and halt SARS-CoV-2 transmission.

**Funding:** São Paulo Research Foundation, Medical Research Council, Wellcome Trust and Royal Society.

## Introduction

Coronavirus disease 2019 (COVID-19) is a severe acute respiratory infection that emerged in early December 2019 in Wuhan, China^1^. COVID-19 is caused by the severe acute respiratory syndrome coronavirus 2 (SARS-CoV-2), an enveloped, single-stranded positive-sense RNA virus that belongs to the *Betacoronavirus* genus, *Coronaviridae* family^2^. SARS-CoV-2 is phylogenetically similar to bat derived SARS-like coronaviruses^3^. Human-to-human transmission occurs primarily via respiratory droplets and direct contact, similar to influenza viruses, SARS-CoV and Middle East Respiratory Syndrome virus (MERS-CoV)^4^.

The most commonly reported clinical symptoms are fever, dry cough, fatigue, dyspnoea, anosmia, ageusia or some combination of these symptoms^1,4-6^. SARS-CoV-2 spread rapidly and as of April 23, 2020, more than 2.7 million cases have been confirmed across the globe, resulting in at least 187,330 deaths^7^.

Brazil identified its first case on February 25, 2020. Despite a prompt public health response, Brazil now accounts for a third of all cases reported in Latin America (46,701 confirmed cases, including 2,940 deaths, as of April 23, 2020)^7^. In this study, we describe the epidemiological, demographical and clinical characteristics from the early phase of the COVID-19 epidemic in Brazil.

## Methods

### Ethical approval

The study was supported by the Brazilian Ministry of Health and ethical approval was provided by the national ethical review board (Comissão Nacional de Ética em Pesquisa, CONEP), protocol number CAAE 30127020.0.0000.0068.

### Individual-level data on notified cases from Brazil

To investigate individual-level diagnostic, demographic, self-reported travel history, place of residence and likely place of infection, differential diagnosis for other respiratory pathogens, as well as clinical details, including comorbidities, we collected case data notified to the REDCap database^8^ from February 25 to March 25, 2020. Data was contributed by public health and private laboratories. Diagnosis and case definitions (see **Appendix, pp**.**1**) were based on World Health Organization (WHO) interim guidance. To explore the time-lag between the number of imported cases and of local cases we used the Granger causality test^9^.

### Geospatial analysis of COVID-19 cases, demographic and socio-economic data

Based on data from the first COVID-19 reports in Brazil^10^, we hypothesized that rates of incidence and testing for COVID-19 are higher in areas of higher per capita income. For the Greater Metropolitan Region of São Paulo (GMRSP), *per capita* income at the GMRSP neighbourhood level (517 zones) were retrieved from the 2017 *Pesquisa Origem e Destino* survey (www.metro.sp.gov.br/pesquisa-od/). 13,913 notified cases (COVID-19 confirmed, ruled out, and without final diagnosis) resident in the GMRSP were geocoded based on self-reported address using the Galileo algorithm and verified using Google API. *Per capita* income for each zone was linked to each notified case based on residential address. We compare *per capita* income for all notified cases between those tested (positive and negative) and untested, and for confirmed cases by RT-PCR. Full details on the statistical analysis can be found in the **Appendix, pp**.**1**.

### Basic reproduction number (R0) estimation

To quantify transmission potential of COVID-19 in Brazil, an exponential model was used to represent the incidence of COVID-19 at the national level and in São Paulo and Rio de Janeiro states. Time series of confirmed cases were modelled as samples from a negative binomial distribution with a mean equal to a fixed portion of the incidence. The analysis was carried out in a Bayesian framework with uninformative priors on all parameters apart from the removal rate, which was given an informative prior. The informative prior ensured that the average duration for which an individual is infectious is 5 to 14 days^11^ (**Appendix, Figs. S1-S2**). Standard diagnostics were used to check whether the Markov Chain Monte Carlo (MCMC) samples were satisfactory. Full details of the model used, the estimation process and convergence of Markov Chain Monte Carlo chains can be found in the **Appendix, pp**.**2**.

### Univariate and multivariate analysis

To investigate which factors are associated with a confirmed COVID-19 result and with hospitalization summary statistics were calculated for continuous variables and for categorical variables and summarized as medians (range and interquartile range, IQR), as appropriate. Missing data were removed (assumed missing at random) (see **Appendix Table S1 and Fig. S3**). Uni- and multivariate analysis included only cases with complete information for the relevant variables. These analyses compared demographics, symptoms, clinical signs and comorbidities between confirmed COVID-19 cases (RT-PCR positive) and ruled-out COVID-19 cases (RT-PCR negative). Additionally, separate multivariate logistic regression models were built to predict hospitalisation (binary variable: hospitalised vs. not hospitalised) based on symptoms, clinical signals and comorbidities, and to predict testing status (positive or negative for RT-PCR SARS-CoV-2). The associations between the outcome and independent variables were reported as adjusted odds ratios (AOR) with 95% confidence intervals and likelihood ratio test (LRT) using the univariate and multivariate logistic regression models. Model diagnostics were performed to check for model specification errors, multicollinearity and influential observations. A 0·05 significance level was applied.

## Results

By March 25, 2020, four weeks after the first report of COVID-19 in Brazil, 67,344 COVID-19 cases had been notified as COVID-19 suspected infections from 172 cities across all five administrative regions of Brazil. Of these, 1,468 cases were confirmed (2·18% of all notified cases) and notified through the REDCap system (**Fig. 1A**), including 1,144 cases (77.9% of 1,468) diagnosed by RT-PCR and 324 (22.1% of 1,468) on clinically epidemiological grounds. During this period, an additional 965 aggregated cases were notified to the Ministry of Health, totalling 2,433 confirmed cases during the first month. Based on notifications via REDCap, 35% (517 of 1,468) of confirmed cases were imported. Of these 326 had the country of travel recorded. The USA and Italy accounted for the majority of the reported imported cases (USA: 82 [25·2%] and Italy 71 [21·7%] of 326 imported cases) (**Fig. 1B**). The epidemic curves of locally-acquired cases followed the curves from imported cases with a lag of two days (Granger causality test) (**Fig. 1A**).

**Figure 1.**
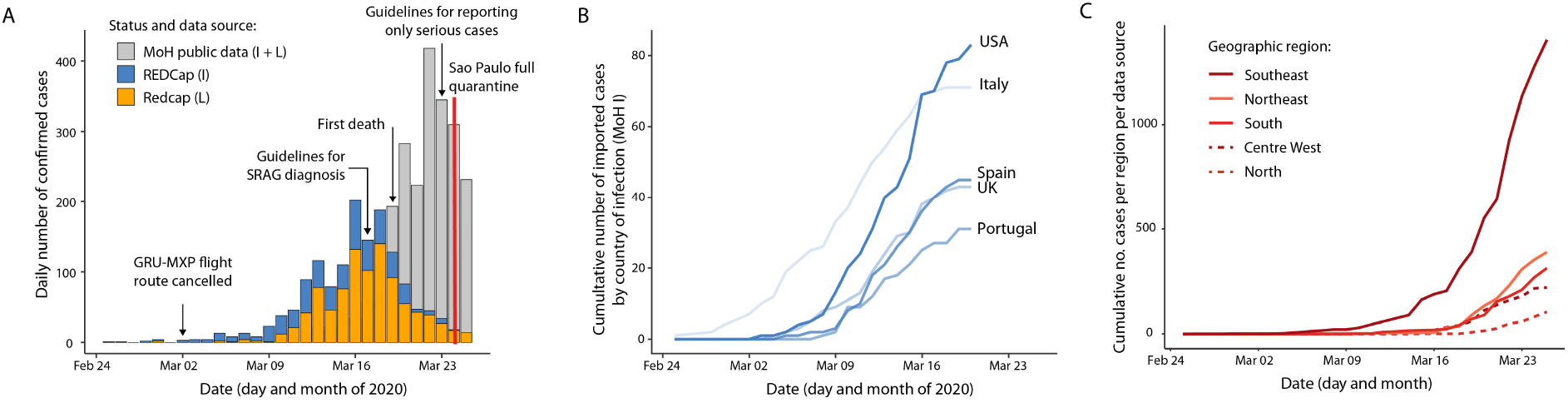
Epidemiological and demographic characteristics of the first confirmed COVID-19 cases within the first 4 weeks of the epidemic in Brazil. **A**. Number of confirmed cases in travellers (blue) and local transmission (orange) from REDCap database. Grey bars show number of aggregated cases reported to Ministry of Health (covid.saude.gov.br/). **B**. Imported cases by self-reported country of infection from REDCap database. **C**. Aggregated cases reported to Ministry of Health by geographic region. GRU = Guarulhos São Paulo International Airport, MXP = Malpensa Milan International Airport, MoH = Ministry of Health. L = Local, I = Imported, and SRAG = Severe acute respiratory syndrome.

According to aggregated data, most confirmed cases were reported in São Paulo (862 [35·4%] of 2,433) followed by Rio de Janeiro (370 [15·2%]). To estimate the basic reproduction number (R_0_) in these locations used a Bayesian approach to fit an exponential growth model to COVID-19 aggregated incidence data. Consistent with previous studies in China and overseas^12^, we find that epidemic spread in São Paulo and Rio de Janeiro states is characterized by similar R_0_ values of 2·9 (95% CI 2·1-4·4) and 2·9 (95% CI 2·2-4·5). The R0 for Brazil was slightly higher with median of 3·2 (95% CI 2·4-5·4) (**Fig. 2**).

**Figure 2.**
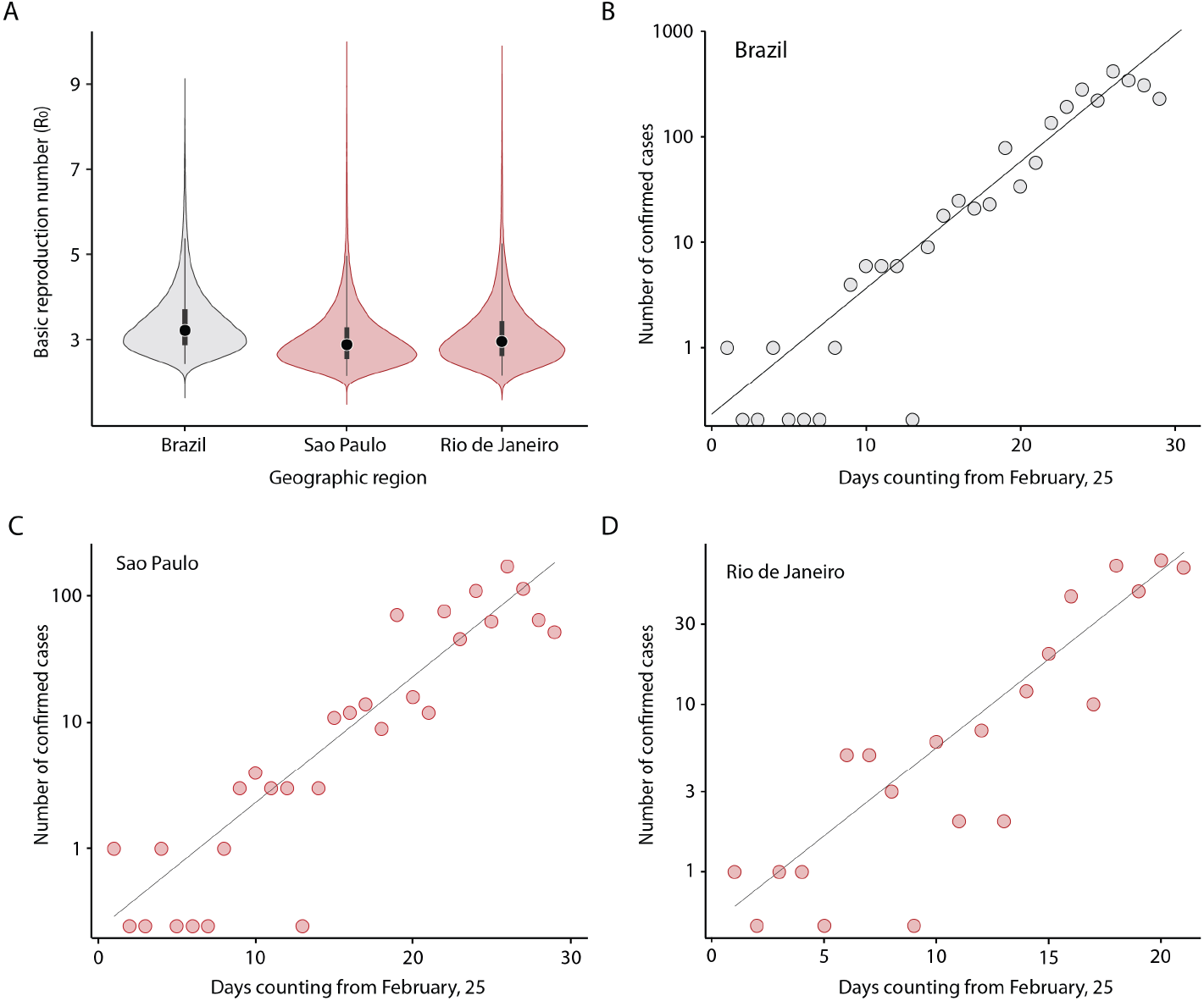
R0 in early phase of the COVID-19 epidemic in Brazil. **A**. Violin plots showing the R0 for COVID-19 estimated using the MoH public data by the March 25, 2020 (n=2,433 cases). **B**. Model fit from point estimate to confirmed cases across all of Brazil, **C**. São Paulo state, **D**. Rio de Janeiro state (see Appendix for details).

Analysis of the age-sex structure of confirmed and notified cases compared to the Brazilian demographic structure revealed a disproportionately lower proportion of confirmed COVID-19 infections reported in younger categories (0**–**9, 10–19 years of age) and a slightly higher proportion in middle-age categories (20–29 and 30–39 years of age) (**Fig. 3**). Specifically, compared to the proportion of the total Brazilian population per age category, the proportion of confirmed COVID-19 infections in the 0**–**9 and 10–19 years of age categories are 16·4- and 5·3-fold lower compared to Brazilian demographic structure (**Fig. 3A**).

**Figure 3.**
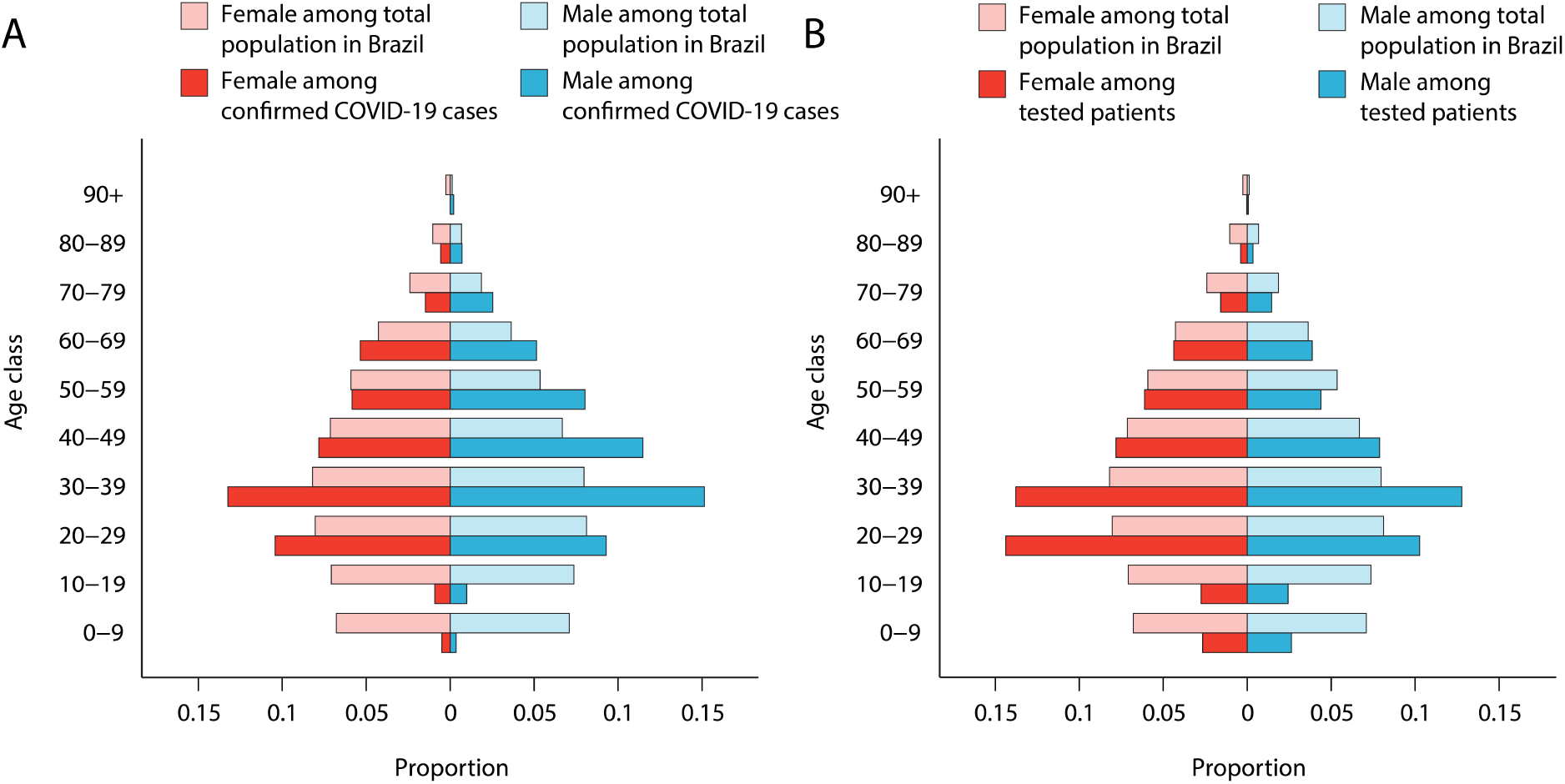
Demographic profile of confirmed COVID-19 (A) and notified (B) cases and total population in Brazil. Age classes are shown on the left. Proportion of confirmed COVID19 and notified cases from the REDCap database for each age-class category are shown as filled bars. Proportion (%) of the country’s population in each age-sex class is shown as faded bars.

We found that most confirmed cases were in males (776 [54·7%] of 1,420 – 46 confirmed cases had missing information for sex and/or age) (**Fig. 3A**). The median age of cases was 39 years (IQR, 30**–**53, range: newborn**–**93 years). Nearly half (695 [48·9%] of 1,420) of the confirmed cases were in the age range of 20 to 39 years of age (**Fig. 3A**). Similarly, 51·6% (2,288 of 4,438) of cases tested for SARS-CoV-2 belonged to this age-group (**Fig. 3B**), which is substantially higher than the corresponding fraction of the Brazilian population (68,451,093 [32%] of 211,755,692). 9·5% (133) of cases were health care workers. Overall, only four newborns, three infants (6 to 8 month-old), ten children (1 to 12 years old), and twelve adolescents (12 to 17 years old) were diagnosed with COVID-19. In addition, nine patients were pregnant, one in the first trimester, one in the second trimester, four in the third trimester and 3 had missing information). Six cases were HIV-positive.

Nearly a third of confirmed cases (462 [36·2%] of 1,277 - 191 confirmed cases had missing information for contact with confirmed case) reported close contact with another confirmed case. After categorizing exposure in travel (international), home (household), work (including schools), and health facility (healthcare workers), we found that over half reported having had contact with a suspected case at their workplace (120 [33·2%] of 385) or at home (88 [22·9%] of 385) (**Fig. 4A**).

**Figure 4.**
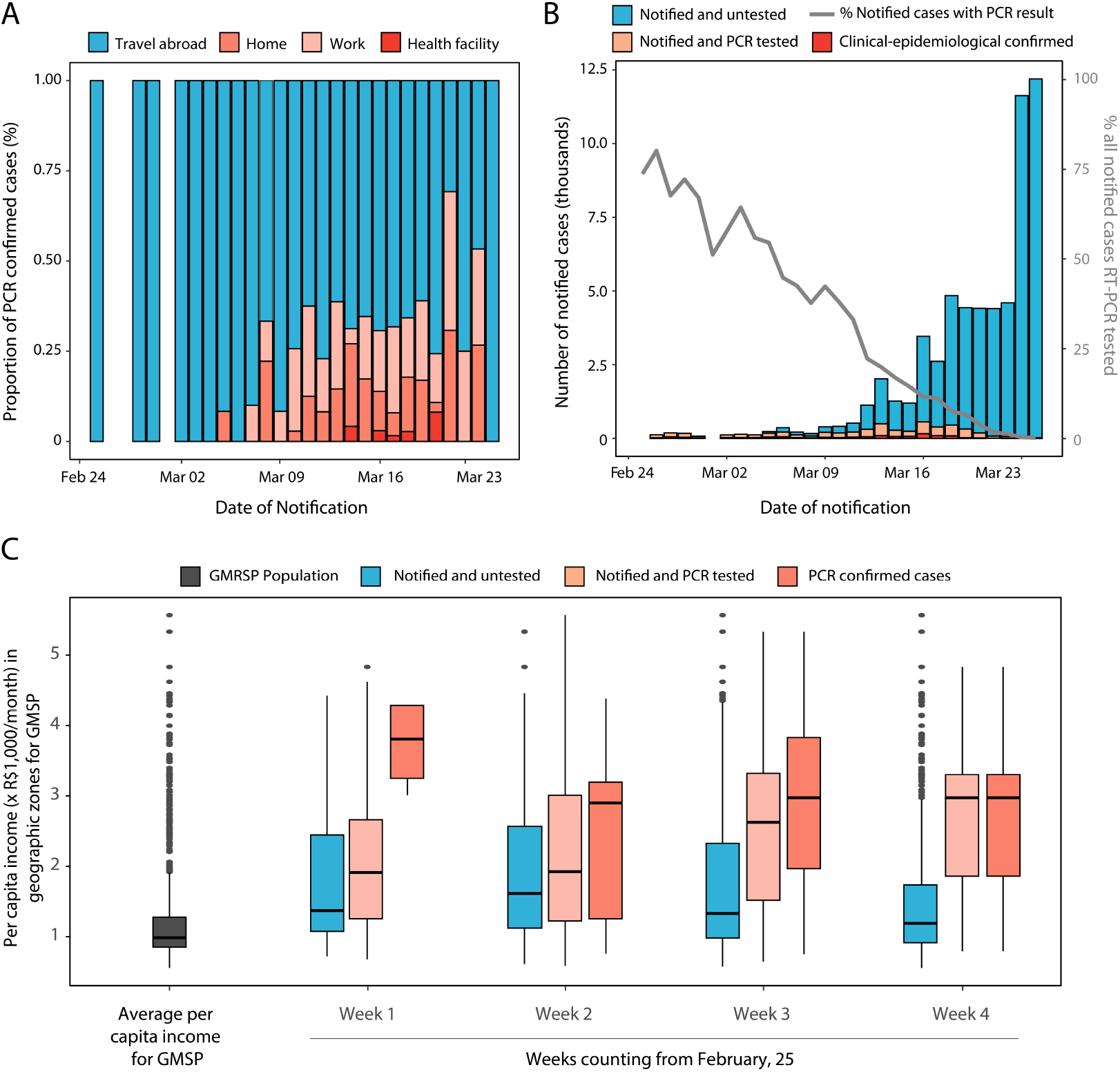
COVID-19 diagnosis and socioeconomic factors in the GMRSP. **A**. Self-reported source of exposure. **B**. Total number of cases notified according to classification status from February 25 through March 25, 2020 (bars), and the proportion of notified cases being tested (line). **C**. Distribution of per capita income based on neighbourhood of residence for all notified COVID-19 cases grouped according to testing status (tested vs. untested), and for RT-PCR confirmed, in the early-phase of the epidemic. The overall distribution of average *per capita* income for 517 zones in the GMRSP weighted by population size is shown on the left of panel **3C**.

**Figure 4B** shows changes in notified confirmed and notified untested COVID-19 cases in Brazil over the study period. Two thirds (586 [66·9%] of 876) of diagnostic tests were performed in private medical laboratories where costs varied typically between 300-690 Brazilian Reais (BRL) (for context, current minimum monthly salary is 1,045 BRL).

To test whether notified tested cases were associated with socioeconomic status, we evaluated the association between COVID-19 diagnosis and socioeconomic status in the subset of cases in the Greater Metropolitan Region of São Paulo (GMRSP) region with geocoded residential information using an ordinal probit model. We found that the proportion of tested cases in GMRSP increased as income per capita increases (z-score = 0.19, likelihood ratio test *P*-value <0.01) (**Fig. 4C, Table S2**). Moreover, the increase in the proportion of tested cases for a unit-increase in income is higher in weeks 2, 3 and 4 compared to week 1. For the range of income per capita observed, given the same amount of income per capita, the proportions of tested cases were lower in weeks 2, 3 and 4 than week 1. Overall, there was a noticeable upwards trend in the association between testing rate and per capita income uncovering a widening socioeconomic disparity in testing practice as the number of cases expands. The income distribution of the untested fraction increasingly approximates the average for GMSP, whereas the tested and confirmed cases (both laboratory and clinical epidemiological) are consistently higher over the study period.

We also analysed the results for other respiratory pathogens tested in Brazil as part of the differential diagnosis by Central Public Health Laboratories and National Influenza Centres (Brazilian Ministry of Health). Respiratory viruses most frequently identified in patients with suspected, but negative diagnosis of COVID-19 were influenza A virus (347 [14·3%] of 2,429), influenza B virus (251 [10·3%] of 2,429) and human rhinovirus (136 [5·6%] of 2,429). We found co-detection of SARS-CoV-2 with six other respiratory viruses, the most frequently were with influenza A (11 [0·5%] of 2,429) and human rhinovirus (6 [0·2%] of 2,429) (**Appendix, Fig. S4**).

We next analysed most common symptoms for confirmed COVID-19 cases in Brazil and which symptoms were linked to hospitalization. Most patients with confirmed COVID-19 (1,351 [92%] of 1,468) reported at least one symptom, the most common being cough (1,040 [70·8%] of 1,468) and fever (982 [66·9%] of 1,468). Other frequent symptoms reported were coryza (495 of [33·7%] 1,468), sore throat (483 [32·9%] of 1,468) and myalgia (450 [30·7%] of 1,468). The characteristics, symptoms and clinical signs of COVID-19 cases confirmed by RT-PCR positive, ruled out by RT-PCR, and notified COVID-19 cases, but no tested are summarised in **Appendix Table S3**.

In a univariate analysis of4,387 cases with a final classification as confirmed (n=1,101) or discarded (COVID-19 ruled out) (n= 3,286), we found that increasing age, symptoms (cough, difficulty breathing, dyspnoea/tachypnea, sputum production, nasal congestion, nasal flaring, nausea/vomiting, headache, irritability/confusion, difficulty swallowing, intercostal retraction and Alteration on chest auscultation) and clinical signs (fever and conjunctival congestion) were higher associated with a negative SARS-CoV-2 results (see **Appendix Table S4)**. Overall, a total of 12·5% (184/1,468) of confirmed COVID-19 cases had at least one comorbidity. Most common comorbidities were heart disease, hypertension, diabetes, and chronic respiratory disease.

By March 25, 10% (147 of 1,468) of patients with COVID-19 had been hospitalized, of whom 15·6% (23 of 147) required mechanical ventilation. The date of hospitalization was available for 140 patients. The median time from symptom onset to hospital admission was four days (IQR= 2 to 6 days, range 0 to 26 days). The most frequent symptoms in hospitalized patients with COVID-19 were fever (122 [83%] of 147) and cough (118 [80·3%] of 147). Multivariable logistic regression showed that chest X-ray abnormalities (AOR: 55·1 [18.88-121.24], p<0·001) and O2 saturation <95% (AOR: 14·80 [4·33-55], p<0·001) were strongly associated with hospitalization (**Figure 5A**). Most hospitalized patients were male (87 [59·2%] of 147). The median age of hospitalized COVID-19 cases was 55 years of age (IQR=40-68), ranging from newborn to 93 years of age; 24·25% (36 of 147) of the hospitalized cases were aged ≤39 years. One of the four newborns was hospitalized with fever, cough, dyspnoea/tachypnoea, altered chest radiology, and abnormal findings on auscultation, but did not require mechanical ventilation. Also, one out of six HIV patients that were diagnosed with COVID-19 was hospitalized and underwent mechanical ventilation. None of the reported cases in babies, children, or pregnant women required hospitalization.

**Figure 5.**
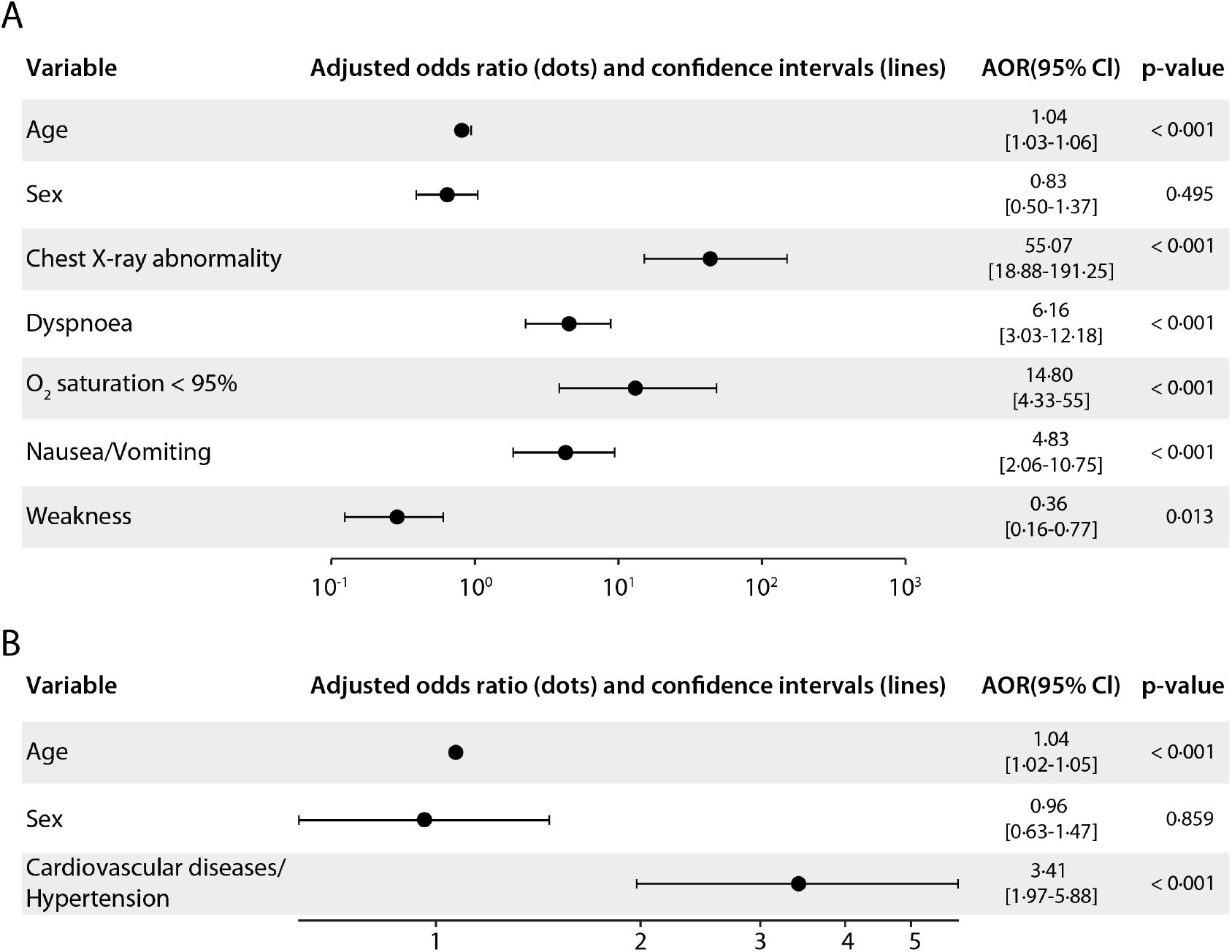
Multivariable logistic regression predicting hospitalisation among RT-PCR SARS-CoV-2 positive cases. **A**. Analysis of symptoms and clinical signs. **B**. Analysis of reported comorbidities.

A large proportion (59 [40·1%] of 147) of hospitalized patients had at least one comorbidity, and the most common were cardiovascular disease/hypertension (47 [29·9%] of 147), diabetes (14 [9·5%] of 147), other respiratory diseases (11 [7·5%] of 147) and solid or haematological neoplasm (7 [4·8%] of 147). Proportions in general population for cardiovascular disease and diabetes are respectively 4·2%, and 6·2%^13^. Multivariable logistic regression analysis showed increasing odds of hospitalization in patients with cardiovascular diseases/hypertension (AOR: 3·41 [1·97-5·87], p<0·001) (**Figure 5B**). Interestingly, age was not significantly associated with hospitalization after accounting for co-morbidities.

Four deaths due to COVID-19 were recorded, and 48 patients remained hospitalized during the study period. Based on the available discharge dates, 3·5% (51 of 1,468) confirmed cases had recovered from the COVID-19 infection by March 25, 2020. The median age among the four deaths was 60 years (IQR = 56 **-** 66), ranging from 49 to 74-years of age) and with a sex ratio of 1:1. The median time from the symptom onset to death was seven days (IQR = 4 **-** 9·5, range, 3 to 14 days). Of the four fatal cases, one had cardiovascular disease/hypertension, one had both cardiovascular disease/hypertension and renal disease, and two fatal cases had no reported comorbidities. Only one case had reported close contact with a confirmed COVID-19 case reinforcing that local transmission was already well established in Brazil by March 25, 2020.

## Discussion

These findings provide evidence that SARS-CoV-2 transmission in Brazil shifted rapidly from a scenario of imported to local transmission. We found that the proportion of tested cases is higher in zones with higher *per capita* income. We showed that during the first month of COVID-19 in Brazil, only 33·1% of the reported confirmed cases were conducted in public health laboratories. Our results support similar transmission potential (R0) of SARS-CoV-2 in Brazil to other geographic regions. Overall, our clinical findings demonstrate that chest X-ray abnormalities and O2 saturation <95% are strongly associated with hospitalization. The combination of universal access to diagnostic and the success of interventions will dictate the fate of COVID-19 in Brazil. Overall, these findings filled in a gap in our understanding of COVID-19 early establishment in Latin America.

We identified several limitations in our study. First, detailed individual-level data was only available for the first month of the epidemic in Brazil. Moreover, several cases had incomplete documentation, such as hospitalization date, mechanical ventilation, and travel history. Real-time aggregated data and open-access open line lists have the potential to provide real-time insights into transmissibility^14^. Secondly, our retrospective study has focused predominantly on symptomatic patients (92%) that presented themselves to health services for testing. Therefore, we cannot describe the full spectrum of disease. Population-based serologic surveys are urgently needed to properly determine the asymptomatic and oligosymptomatic fraction. Finally, many patients remained hospitalized when the dataset was extracted, and, we were unable to estimate clinical outcomes given the long duration of infection.

Together with changes in surveillance guidelines, socioeconomic bias in testing suggests that the number of confirmed case counts may substantially underestimate the true number of cases in the population. Additional reasons for underreporting include (i) a significant proportion of asymptomatic infections^15^, (ii) people with mild and even moderate disease are unlikely to present to health services for testing, (iii) limited testing capacity in public health service in Brazil in face of the large number of cases due to delays in importing reagents and kits used in molecular testing. Close monitoring of state- and municipality-level data will further help to inform mitigation strategies.

Our results suggest that approximately 50% of the COVID-19 cases in Brazil were skewed towards age groups between 20 to 39 years with substantially fewer cases in younger age groups. This pattern could be explained by (i) a higher risk of exposure of this group due to more frequent international travel (travel bans were only implemented on March 23, 2020), and (ii) younger age groups being less likely to acquire an infection and/or less likely to acquire significant symptoms upon being infected^16^.

COVID-19 infections were reported in paediatric and pregnant patients^17-19^. Paediatric infection appears to typically be of mild or moderate severity; we observed a similar proportion of asymptomatic infections compared to reports in 36 children in China (24% vs. 28%)^19^. Also, the onset symptoms of pregnant women were similar to those reported in non-pregnant adults with COVID-19 infection. On the other hand, proportion of hospitalisation of paediatric patients in Brazil was lower than those observed for children in China (3.3% vs. 38.9%)^19^. Also, in contrast to China, none of the pregnant women that tested positive for COVID-19 in Brazil had pneumonia or were hospitalized^17,18^. However, the absence/lower number of hospitalisations could be explained by resource availability and local clinical practice guidelines. Despite the small sample size, our findings in pregnant and paediatric patients in the early-phase COVID-19 pandemic in Brazil require further understanding of SARS-CoV-2 infection in these groups.

Although clinical features in Brazil are similar to those recently reported in other countries ^1,4,5^, we observed that 8% of confirmed cases reported no symptoms. This should not be considered as an estimate of the asymptomatic fraction. Firstly, it is not possible to distinguish true asymptomatic infections from cases in the pre-symptomatic phase. Secondly, routinely collected data tends to be incomplete. Thirdly, these cases were tested because they were in contact with a known confirmed case. Lastly, there is an ascertainment bias towards symptomatic infections due to the case definition used for notification (Appendix). Other estimates of the asymptomatic fraction have varied widely, including 18% on the Diamond Princess ship^15^, 50-75% in the Italian village of Vo’Euganeo^20^ and 31% based on repatriation flight screening^21^.

Overall, 10% of COVID-19 cases in Brazil were hospitalized compared to 19% in the USA^22^. As mentioned above, these differences may reflect factors other than disease severity, for example, resource availability, local clinical practice guidelines and testing availability. On the other hand, they may also reflect right censoring, whereby cases that were notified towards the end of the period studied had not yet been hospitalized. This would be expected given the median lag of four days between symptom onset and hospitalization observed in Brazil. Although age was not a risk factor for hospitalization after controlling for comorbidities, is should be noted that the age distribution among patients who were hospitalized differed from that reported in China, with a higher proportion of younger (<39 years: Brazil, 24.5% vs. China, 10%) and older patients (>70 years: Brazil, 21.8% vs. China, 15%)^18^. However, such comparisons need to be taken cautiously due to different testing and notification practises in the two countries.

We showed that patients with pre-existing cardiovascular diseases/hypertension were at increased risk of hospitalization. The prevalence of at least one comorbid condition among infected individuals in Brazil was similar to that reported in China (12.5% *vs*. 10.5%)^23^. Previous studies suggest that persons with underlying health conditions, such as cardiovascular, diabetes and chronic lung diseases, appear to be at higher risk for severe COVID-19 infection than persons without these conditions^22,24^. Pre-existing cardiovascular disease appears to be particularly important, potentially due to the involvement of the renin angiotensin system signalling pathway^25^.

This study provides new information on co-circulation and co-detection of other respiratory pathogens in the early phase of the COVID-19 epidemic in Brazil. Particularly, we found co-circulation of eight other respiratory viruses, the most common respiratory infections were influenza A and B, and human rhinovirus (HRV). Co-detection of SARS-CoV-2 with influenza A and human metapneumovirus (hMPV) have also been reported in China^26,27^. Here we found co-detection of SARS-CoV-2 with influenza A and hMPV, and we expanded the description of the other multiple co-detection scenarios of SARS-CoV-2 with other respiratory viruses, including HRV, influenza B, human respiratory syncytial virus, and other coronaviruses (i.e. coronavirus 229E/NL63, hCoV OC43/HKU1). Although, viral co-infection has been reported with many other respiratory viruses, no difference in clinical disease severity between viral co-infection and single infection has been reported^28^.

In conclusion, we provide the first description of COVID-19 in Brazil. Our study provides crucial information for diagnostic screening and health-care planning, and for future studies investigating the impact of non-pharmaceutical and pharmaceutical interventions in mitigating COVID-19 transmission.

## Data Availability

All data mentioned in the manuscript are available.

## Contributors

JC, JPC, LFB, CAP, VHN, WMS, DSC, AEZ, JM, FCSS, PSA, FG, AASS, BG, CHW, RHMP, SL, NG, SBO, VBGP, FG, WAFA, FFSTF, EMM and WKO collected the epidemiological, spatial and clinical data and processed statistical data. NRF, WMS, LFB, CHW, JPC, DCS, JM, ECS, PM, SL, RHMP, LA, AASS, GL, AT, MFVG, MUGK, RSA, NA, PM, OJB, IOMS, NG, GL, OGP, AEZ, and JC interpreted the results and wrote the manuscript. All authors read and revised the final manuscript. JC, WMS, LFB, MFVG, and NGJ are responsible for summarising epidemiological and clinical data.

## Acknowledgments

The authors thank the clinicians and epidemiologist for technical support. This work was supported by a FAPESP (2018/14389-0) and Medical Research Council and CADDE partnership award (MR/S0195/1). NRF is supported by a Sir Henry Dale Fellowship (204311/Z/16/Z). WMS is supported by the São Paulo Research Foundation, Brazil (No. 2017/13981-0). OJB was funded by a Sir Henry Wellcome Fellowship funded by the Wellcome Trust (206471/Z/17/Z). VHN and CAP were supported by FAPESP (2018/12579-7). AEZ and BG are supported by Oxford Martin School.

